# Diagnostic performance of Attenuated Total Reflectance Fourier Transform Infrared (ATR-FTIR) spectroscopy for detecting osteoarthritis and rheumatoid arthritis from blood serum

**DOI:** 10.1101/2024.10.11.24315302

**Authors:** Minna Mannerkorpi, Shuvashis Das Gupta, Lassi Rieppo, Simo Saarakkala

## Abstract

Osteoarthritis (OA) and rheumatoid arthritis (RA) are the two most common rheumatic diseases worldwide, causing pain and disability. Both conditions are highly heterogeneous, and their onset occurs insidiously with non-specific symptoms, so they are not always distinguishable from other arthritis during the initial stages. This makes early diagnosis difficult and resource-demanding in clinical environments. Here, we estimated its diagnostic performance in classifying ATR-FTIR spectra obtained from serum samples from OA patients, RA patients, and healthy controls. Altogether, 334 serum samples were obtained from 100 OA patients, 134 RA patients, and 100 healthy controls. The infrared spectral acquisition was performed on air-dried 1µl of serum with a diamond-ATR-FTIR spectrometer. Machine learning models combining Partial Least Squares Discriminant Analysis (PLS-DA) and Support Vector Machine (SVM) were trained to binary classify preprocessed ATR-FTIR spectra as follows: controls vs. OA, controls vs. RA, and OA vs. RA. For a separated test dataset and the validation dataset, the overall model performance was better in classifying OA and RA patients, followed by the RA and controls, and lastly, between OA and controls, with corresponding AUC-ROC values: 0.84, 0.76, 0.72 (test dataset) and 0.94, 0.92, 79 (validation dataset). In conclusion, this study reports robust binary classifier models to differentiate blood serum from the two most common rheumatic diseases, showing the potential of ATR-FTIR as an effective aid in rheumatic disease classification.

## 1. Introduction

Osteoarthritis (OA) and rheumatoid arthritis (RA) are the two most common forms of arthritis, responsible for disability and chronic pain in adults, causing significant socioeconomic burden worldwide [1,2]. Both diseases are highly heterogeneous, with a complex pathophysiology of an unknown origin. OA is best characterized as a degenerative whole-joint disease most frequently occurring in knees, where various joint tissues undergo structural modifications, leading to failure. [3,4]. RA is a systemic inflammatory autoimmune disease often characterized as symmetrical polyarthritis, affecting small and large synovial joints throughout the body. In addition, some patients develop extra-articular disease manifestations, such as interstitial lung disease, vasculitis, neuropathy, serositis, glomerulonephritis, inflammatory eye involvement, felty syndrome, myopathy, and amyloidosis. [5,6] It is well-recognized that in both OA and RA, the disruption at the molecular level occurs years before the clinical symptoms, making them asymptomatic and thus difficult to diagnose in the initial stages. [5,7] Therefore, the diagnostic process for OA and RA is complex, relying on a combination of clinical exams, serological tests, and imaging, as there is no single diagnostic marker. Current classification criteria (ACR/EULAR [8] and NICE [9]) show varying sensitivity and specificity over time [10,11], highlighting the need for faster, more efficient diagnostic tools for early-stage detection and personalized treatment.

In recent years, Attenuated Total Reflectance Fourier Transform Infrared spectroscopy (ATR-FTIR) studies with blood serum and machine learning (ML) based analysis techniques have been the subject of active research to develop diagnostic methods for various diseases [12–15]. ATR-FTIR is a non-expensive vibrational spectroscopy method that provides a label-free, analytical way to study the biochemical composition of small sample volumes with minimal preparation. In ATR-FTIR spectroscopy, the sample is placed in contact with the ATR reflection element that directs the IR radiation into the sample. Due to the optical properties of the ATR element, the IR light travels through the element as an evanescent wave as a frequency-dependent absorption occurs within the sample if the chemical bond of the molecule is vibrating at the same frequency as the incident IR radiation. [16] The absorbance spectrum represents the convolution of individual spectra initiated by all covalent bonds of infrared-active biomolecules, providing characteristic information of the biochemical composition of the sample, i.e., the molecular “fingerprint.”

This is also evidenced by previous studies demonstrating the potential of ATR-FTIR and FTIR spectroscopy to discriminate rheumatic diseases from human blood serum samples [17–22] and synovial fluid [23]. Although synovial fluid can reflect a more detailed metabolic status of OA and RA and may lead to better diagnostic results, as the early study suggests, the clinical value of blood serum is higher since it is more available and better represents the systematic effects of a disease in individuals, considering the complexity of rheumatic diseases. [24,25] Recent studies in food science [26], pharmacology [27], and cancer research [28–31] have utilized Partial Least Squares Discriminant Analysis (PLS-DA) as a dimension reduction technique in combination with Support Vector Machine (SVM) to classify spectral data.

In this study, we assessed the potential of ATR-FTIR spectroscopy in diagnosing two common rheumatic diseases. We aimed to investigate the classification of OA and RA patients from healthy controls using blood serum with IR spectra using an innovative approach, combining both PLS-DA and SVM algorithms. We determined the robustness and potential of the model in diagnosing arthritic diseases.

## 2. Material and Methods

### 2.1 Ethics and Study Design

All the blood serum samples were collected from a local biobank, Biobank Borealis of Northern Finland (Oulu, Finland), governed by the Finnish Biobank Act 688/2012. The biobank approved study approval requests with a detailed study protocol and a sample request (corresponding biobank project number: BB_2021_5009) and did not require a statement from the local ethics committee. Table 1 represents the available inclusion and exclusion criteria for the participants of this study.

**Table 1.** The inclusion and exclusion criteria of the participants.

| Group | Inclusion | Exclusion |
| --- | --- | --- |
| Control | - When none of the exclusion criteria is satisfied | - Tumors<br>- Dyscrasias and immunomechanical disorders,<br>- Metabolic and endocrine diseases<br>- Psoriatic arthritis,<br>- Systemic lupus erythematosus,<br>- Sjögren's syndrome<br>- Crohn's disease,<br>- Ulcerative colitis |
| OA | - Clinically diagnosed knee OA<br>- When none of the exclusion criteria is satisfied |  |
| RA | - Clinically diagnosed seropositive RA<br>- Clinically diagnosed seronegative RA<br>- When none of the exclusion criteria is satisfied |  |

### 2.2 Sample set

The sample set consisted of 334 serum samples from healthy (control group), OA patients, and RA patients. The samples were collected into serum gel sample tubes, following a 30-minute stand time before centrifugation. The centrifugation was performed at room temperature according to the instructions by Nord Lab (https://www.nordlab.fi/) (2500g, 10 min) and stored at −80°C until pipetting. Subsequently, 20µl of serum was sectioned into Eppendorf tubes and then frozen at −80°C until the ATR-FTIR measurements. Table 2 represents the available demographics of the study participants obtained from Biobank Borealis.

**Table 2.** The available demographics of the study participants. The age at the diagnosis is determined based on the time when the first entry of the given diagnosis in the patient information system is found as the principal diagnosis.

|  | RA |  |  |  |  |  | OA |  |  | Control |  |  |
| --- | --- | --- | --- | --- | --- | --- | --- | --- | --- | --- | --- | --- |
|  | Male<br>(n=37) |  | Female<br>(n=97) |  | All<br>(n=134) |  | Male<br>(n=48) | Female<br>(n=52) | All<br>(n=100) | Male<br>(n=48) | Female<br>(n=52) | All<br>(n=100) |
| The Mean Age at sampling<br>± SD | 52.3<br>±11.6 |  | 54.8<br>±10.3 |  | 54.2<br>±10.7 |  | 60.0<br>±9.4 | 58.3<br>±7.0 | 58.2<br>±8.2 | 52.8<br>± 10.4 | 54.3<br>± 10.8 | 53.8<br>±10.6 |
|  | Seropositive |  |  | Seronegative |  |  |  |  |  |  |  |  |
|  | Male<br>(n=33) | Female<br>(n=67) | All<br>(n=100) | Male<br>(n=4) | Female<br>(n=30) | All<br>(n=34) |  |  |  |  |  |  |
|  | 53.6<br>±11.2 | 55.7<br>±10.0 | 55.1<br>±10.4 | 41.0<br>±9.2 | 53.0<br>±10.7 | 51.2<br>±11.1 |  |  |  |  |  |  |
| The Mean Age at diagnosis<br>± SD | 46.2<br>±10.2 |  | 48.7<br>±9.5 |  | 48.0<br>±9.7 |  | 51.8<br>±8.1 | 52.2<br>±5.5 | 52.0<br>±6.8 | - | - | - |
|  | Seropositive |  |  | Seronegative |  |  |  |  |  |  |  |  |
|  | Male | Female | All | Male | Female | All |  |  |  |  |  |  |
|  | 47.6<br>±9.6 | 48.4<br>±9.3 | 48.2<br>±9.4 | 34.5<br>±6.4 | 49.4<br>±10.1 | 47.6<br>±10.8 |  |  |  |  |  |  |

### 2.3 ATR-FTIR measurements

The measurements were performed with an ATR-FTIR spectrometer (Thermo Scientific Nicolet iS5, Thermo Nicolet Corporation, Madison, WI, USA) with an id7-ATR-diamond accessory. For spectral acquisition, the vendor-provided OMNIC™ software was used. Before each measurement, a background spectrum was collected. One microliter of serum was pipetted onto the ATR crystal, and the spectral acquisition was performed within the MID-IR region (400 - 4000 cm^-1^), averaging 64 repeated scans with a spectral resolution of 4 cm^-1^. During each measurement, 10 IR spectra were collected while the serum sample was drying (approx. 20 min) to monitor water evaporation. Only the final (tenth) spectrum without the interfering water band (900 – 1800 cm^-1^ & 3000 – 3700 cm^-1^) [32] was used for further preprocessing and analysis. To guarantee the reliability of the measurements and to prevent any potential distortions resulting from the device or the surrounding environment, the measurement protocol described above was repeated three times and averaged for each sample. Between repetitions, the ATR crystal was cleaned from serum by erasing it with 70% ethanol, then with Virkon, and finally with 70% ethanol.

### 2.4 Spectral pre-processing

All spectral analyses were performed using MATLAB® (MathWorks R2023b, 9.13.0.2105380, Natick, Massachusetts). While the fingerprint region (800 – 1800 cm^-1^) is commonly chosen for its distinct peak characteristics, we also aim to assess if the entire spectral range contributes to the model development, given the intricate nature of rheumatic diseases. Thus, the spectral data was truncated into two separate regions: 800 – 1800 cm^-1^ and 800 – 3700 cm^-1^, which were further processed similarly. [33] After the data truncation, the offset was corrected for the spectra, followed by vector normalization to minimize the non-biochemical effects on the data. In addition, the first and second derivatives of the spectra were calculated to examine whether the derivative spectral data affects the classification performance and enhances the differences between groups. The derivatives were then truncated and normalized. The derivatives were calculated using the Savitzky-Golay filter, a built-in function provided by MATLAB®, with a polynomial order of 2 and a window length of 9.

### 2.5 Binary classifier model

First, a Principal Component Analysis (PCA) was applied to the preprocessed data, as it is a common unsupervised method for dimension reduction before classification analysis. However, PCA could not find discriminative patterns between groups, as shown in Supplementary Figure S2. Furthermore, statistical tests were assessed to find relevant wavenumbers (t-test with permutation); however, no significant wavenumbers were identified. Therefore, a supervised method for dimension reduction for further classification was seen as the most robust approach.

Three separate binary classification models were trained between the groups: 1) control and OA, 2) control and RA, and 3) OA and RA. The well-established chemometric method, PLS-DA, was combined with SVM for classification. PLS-DA is a powerful classification technique for high-dimensional datasets by projecting them into latent variables (LVs) in which Linear Discriminant Analysis (LDA) is then applied [33]. However, in cases where the differences between groups are minimal, the groups may not be linearly separable in latent variable space, making LDA inadequate. Therefore, combining PLS-DA and SVM may provide superior performance compared to PLS-DA alone. The power of SVMs is that they perform better on data with nonlinear characteristics when kernel functions are used. These functions map the data into higher-dimensional space, where a linear separation plane can be found. Here, we used the Gaussian Radial Basis Function: 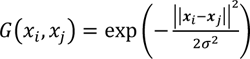 as a kernel.

The training protocol of the PLS-DA-SVM model is summarized in Figure 1. To prevent data leakage and overfitting, the preprocessed data was split into a train set *X*_*train*_ constituting 70% of the data and the rest 30% to test set, *X*_*test*_. Following the division, a MATLAB build-in PLS algorithm was implemented on the training set to construct the PLS-score matrix *t*_*train*_ and weight vector *w*_*train*_. The PLS-score matrix *t*_*train*_, was used as an input in the SVM classifier to train the model.

**Figure 1.**
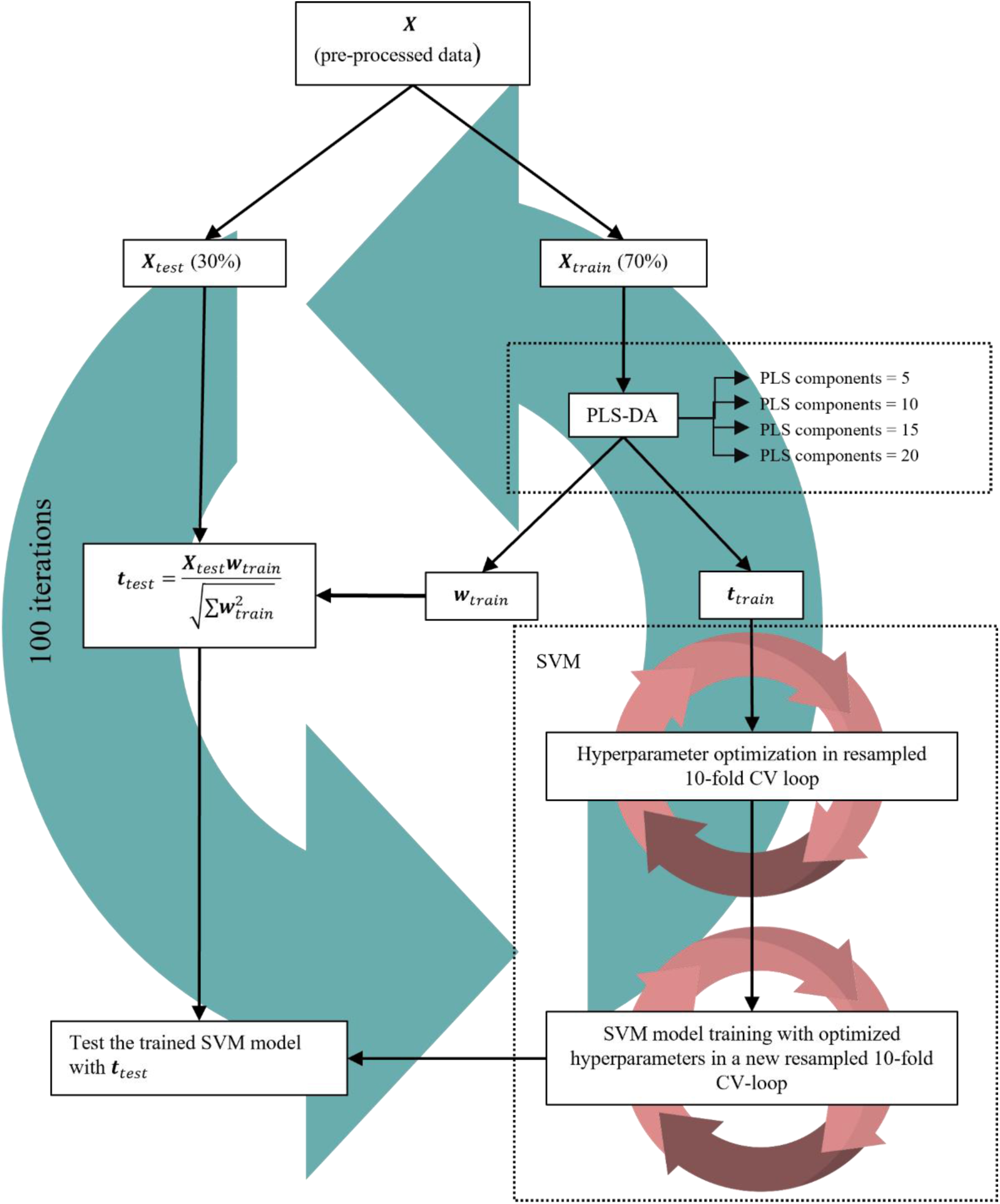
The training protocol for the PLS-DA-SVM model in a nested cross-validation loop. After preprocessing, the spectral data is divided into a train set, *X*_*train*_, and a test set, *X*_*test*_. Following the data division, the PLS-DA algorithm reduces the dimension of *X*_*train*_, and determines the PLS-score matrix *t*_*train*_, and weight vector *w*_*train*_. Subsequently, the *t*_*train*_ is fed to an SVM model training loop. Hyperparameters are optimized in a resampled cross 10-fld cross-validation loop with Bayesian optimization. Then, actual SVM model training with optimized hyperparameters is performed in a new resampled 10-fold CV loop. Finally, the trained SVM model is assessed with *t*_*test*_determined based on the weight matrix calculated from *X*_*train*_.

Three hyperparameters needed to be optimized before the final SVM model training: the number of PLS components for PLS-DA, the Kernel scale σ, and the Penalty coefficient C for SVM. Prior to actual SVM model training, we tried to optimize the number of PLS components by assessing the change in the 10-fold cross-validation (CV) loss of the SVM model as a function of the maximum number of 30 PLS components. However, as illustrated in Supplementary Figure S5, the CV error does not reach a saturation point or reaches zero (indicating overfitting). This makes the PLS component selection based on a cut-off (saturation) point inadequate. Therefore, we trained SVM models with five, ten, fifteen, and twenty PLS components and selected the best one based on the AUC-ROC value of the model. Following the dimension reduction, Bayesian optimization was employed to optimize the kernel scale C and penalty coefficient σ by minimizing (resampled) the 10-fold CV loop.

After the hyperparameter optimization, the SVM model was trained in a new (resampled)10-fold CV loop. After training, the performance of the SVM model was evaluated with *t*_*test*_ determined based on the weight matrix calculated from *X*_*train*_ according to the following formula: 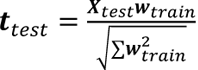

The training protocol described above was iterated 100 times. In each iteration, a new random splitting (resampling) in training and testing sets *X*_*train*_, and *X*_*test*_, was performed, and the results across iterations were aggregated and averaged. This nested CV is recommended for limited data sets, ensuring an unbiased model development and an honest generalization estimate. [34]

## 3. Results

### 3.1 Spectral differences between OA, RA, and healthy patients

Visual examination of the mean spectra reveals minimal biochemical differences between groups, as shown in Figure 2 (all preprocessed spectra are plotted in Figure S1), with assignments of the standard vibrational modes of blood serum. [35] The difference spectrum also reveals minor differences between groups, mainly in locations assigned to Amide I and II peaks and PO2--stretching (Figure 3). Moreover, the first and second mean derivative spectra (supplementary Figure S3-S4) showed no significant differences. Figure 4 represents the data in LV space. The first three LVs do not show linear group separation, thus necessitating a non-linear discrimination method (SVM).

**Figure 2.**
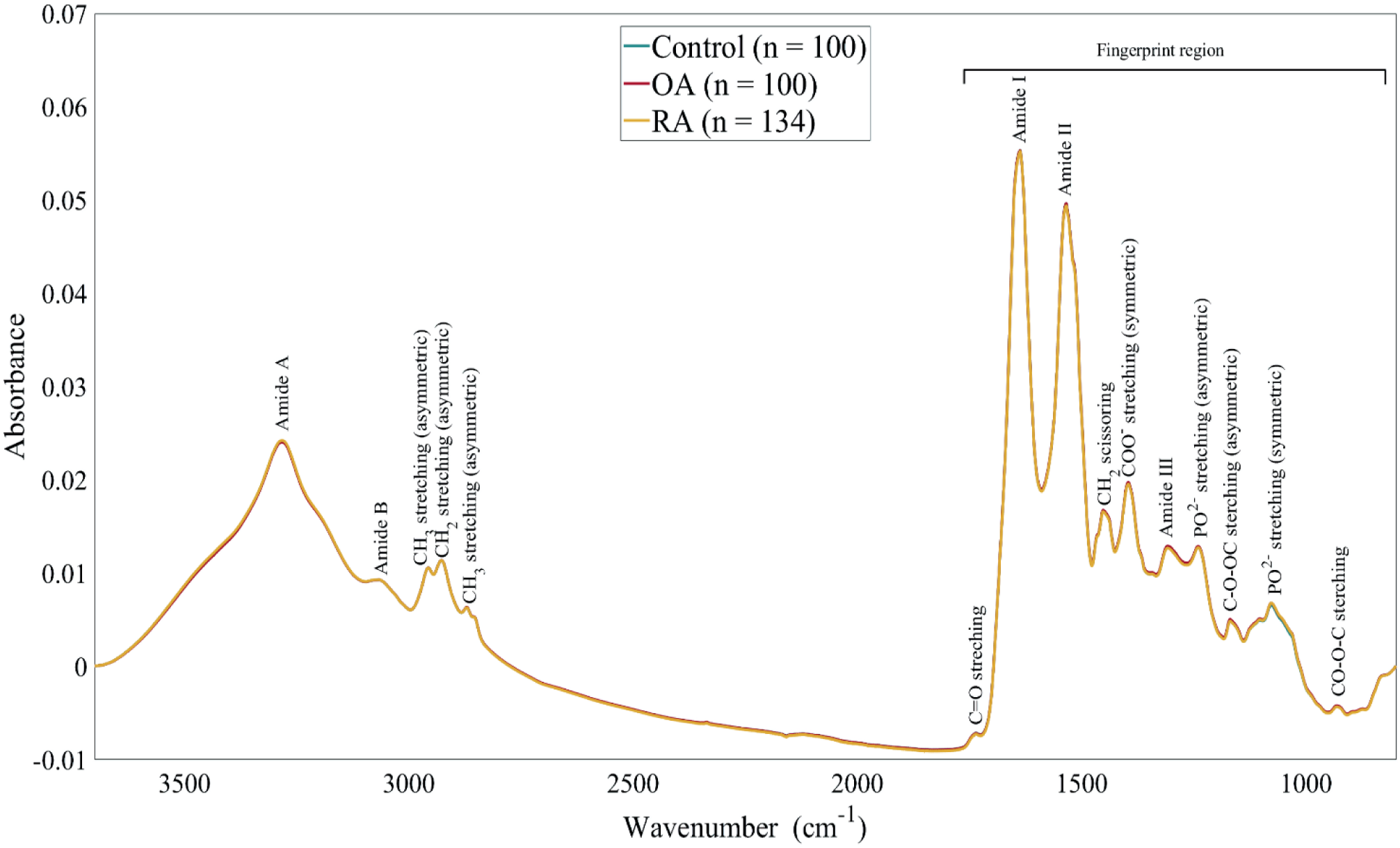
The mean spectra of the three groups in the spectral region of 800 – 3700 cm^-1^, with the fingerprint region (800 – 1800 cm^-1^) highlighted. The blue spectrum represents the mean spectrum of the control group, the red spectrum represents the mean spectrum of the OA group, and the yellow spectrum represents the mean spectrum of the RA group.

**Figure 3.**
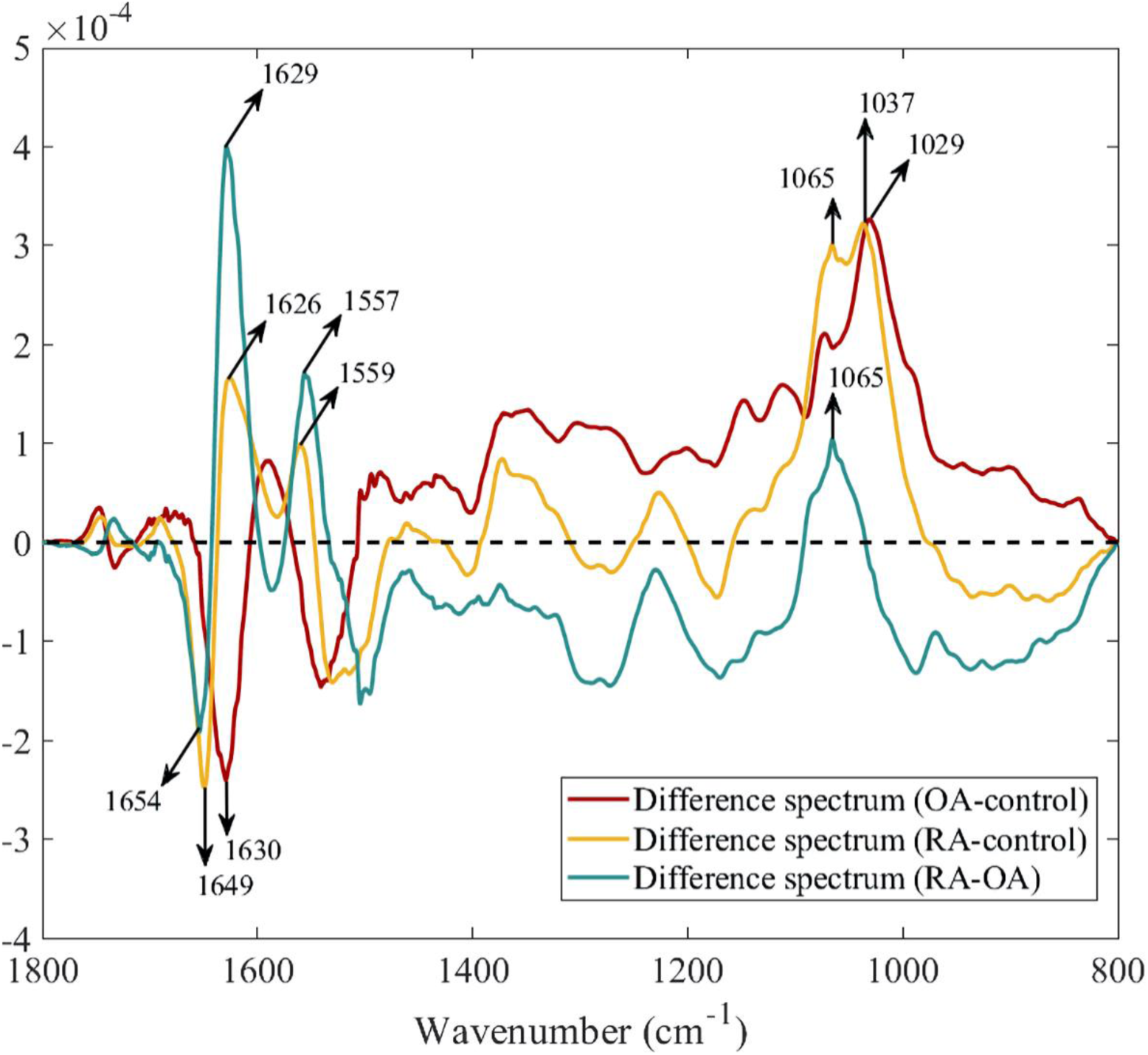
The difference spectra between groups are calculated from the mean spectra represented in Figure 2. The wavenumbers of the most prominent differences are highlighted with a black arrow.

**Figure 4.**
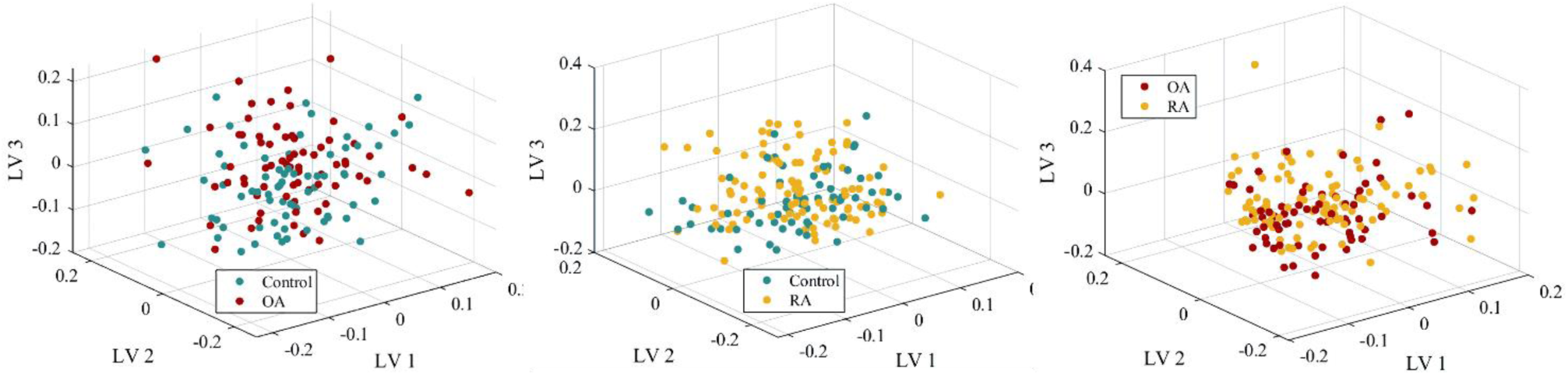
An illustration of normalized spectral training data (800 – 3700 cm^-1^) transformed into latent variable (LV) space. It is evident from the first three LV variables that the linear discrimination method is inadequate and necessitates a non-linear method.

### 3.2 Classification performance

Overall, the PLS-DA-SVM classifier performed best with normalized spectral data at the wavenumber region of 800 – 3700 cm^-1^, the results of which are presented here and summarized in Table 3. The supplementary material (Table S1-S10) presents the classifier’s performance from the fingerprint region of the normalized and derivative data (800 – 1800 cm^-1^).

**Table 3.** The highest-observed performance of the PLS-SVM classifier on validation and test set when normalized spectral data, truncated into the 800 – 3700 cm^-1^ wavenumber region, was used. The averaged performance of PLS-DA-SVM across 100 iterations is represented in the brackets below.

| Performance metrics |  | Number of PLS-components | AUC-ROC Maximum (Average) | Accuracy (%) Maximum (Average) | Sensitivity (%) Maximum (Average) | Specificity (%) Maximum (Average) |
| --- | --- | --- | --- | --- | --- | --- |
| Validation set | Control vs. OA | 10 | 0.79 (0.70) | 73 (66) | 70 (67) | 79 (68) |
|  | Control vs. RA | 20 | 0.92 (0.87) | 84 (81) | 96 (84) | 69 (77) |
|  | OA vs. RA | 20 | 0.94 (0.91) | 89 (85) | 90 (87) | 85 (83) |
| Test set | Control vs. OA | 10 | 0.72 (0.61) | 67 (58) | 70 (62) | 63 (55) |
|  | Control vs. RA | 20 | 0.76 (0.67) | 71 (69) | 72 (76) | 71 (55) |
|  | OA vs. RA | 20 | 0.84 (0.70) | 83 (70) | 78 (76) | 75 (60) |

The PLS-DA-SVM model achieved its best performance when differentiating OA and RA spectra. The best observed AUC-ROC value in the validation set was 0.94, with corresponding accuracy, sensitivity, and specificity of 94%, 89%, and 85%, respectively. The OA vs. RA classification performed best in the validation set with AUC-ROC value, accuracy, sensitivity, and specificity, 0.94, 89%, 90%, and 85%, respectively. The second-best performance was control vs. RA classification (AUC-ROC of 0.92), with accuracy, sensitivity, and specificity of 84%, 96%, and 69%, respectively. However, the classification of OA and control spectra demonstrated a lower AUC-ROC of 0.79, with 73% accuracy, 70% sensitivity, and 79% specificity in the validation set, and 0.72, 67%, 70%, and 63% in the test set. In the test set, OA vs. RA classification maintained the best performance, with an AUC-ROC of 0.84, accuracy of 83%, sensitivity of 78%, and specificity of 75%. The classification between RA and control spectra showed similarly high performance, achieving an AUC-ROC of 0.76, with 71% accuracy, 72% sensitivity, and 71% specificity. However, the OA and control spectra classification demonstrated a lower AUC-ROC of 0.72, with 67% accuracy, 70% sensitivity, and 63% specificity.

In the validation set, the averaged performance across 100 iterations, OA vs. RA classification remained the highest, with an AUC-ROC of 0.91, accuracy of 85%, sensitivity of 87%, and specificity of 83%. The second-best performance occurred in the control vs. RA classification, with an AUC-ROC of 0.87, 81% accuracy, 84% sensitivity, and 77% specificity. The classification of OA and control spectra was the lowest, with an AUC-ROC of 0.70, accuracy of 66%, sensitivity of 67%, and specificity of 68%. In the test set, OA vs. RA classification maintained the best performance, with an AUC-ROC of 0.70, accuracy of 83%, sensitivity of 78%, and specificity of 75%. The second-best performance was control vs. RA classification (AUC-ROC of 0.60), with accuracy, sensitivity, and specificity of 69%, 76%, and 55%, respectively. The OA vs. control classification yielded an AUC-ROC of 0.61, 58% accuracy, 62% sensitivity, and 56% specificity.

## 4. Discussion

In this study, we evaluated the potential of ATR-FTIR spectroscopy to distinguish well-characterized cohorts of OA, RA patients, and a healthy control group based on their blood serum. We trained PLS-DA-SVM hybrid classifier models with ATR-FTIR spectral data and tested the classification performance of different spectral preprocessing approaches and spectral regions of interest. The PLS-DA-SVM models were trained with a nested cross-validation (CV) approach and achieved over 80% AUC-ROC in distinguishing spectra of the RA group from the OA group.

The difference spectrum between the RA and control groups (control mean spectrum subtracted from RA mean spectrum) reveals a positive peak at 1626 cm^-1^, consistent with findings from a previous study that identified the same peak, along with two additional peaks at 1628 cm^-1^ and 1627 cm^-1^, all of which were strongly negatively correlated with RF [22]. We also observed a prominent positive peak in the difference spectrum at 1629 cm^-1^ (OA mean spectrum subtracted from RA mean spectrum), which corresponds well to the two other identified peaks. This wavenumber is associated with the β-sheet of IgG3 and IgG2. [36]. Considering the ATR-FTIR’s sensitivity to conformational changes in proteins and the dominant structure of β-sheet in FC-tail for RF binding, this finding may reflect the conformational differences in the antibody binding sites between individuals. It has recently been reported that these individual conformation patterns can explain the presence of natural RF and pathophysiological RFs in autoimmunity [37] and, therefore, play a role in the difference peaks observed. Moreover, the glycosylation of IgG is linked to both OA and RA, which could have contributed to the subtle differences observed between bands in the carbohydrate region (1180-1000 cm⁻¹), as it is reported to be sensitive to protein glycosylation in human plasma [38,39].

Prior studies have performed classification using the whole spectral range (700 – 4000 cm^-1^) [17] or selecting significant wavenumbers from various ranges (650 – 4000 cm^-1^ and combinations of 450 – 1700 cm^-1^ and 1701 – 4000 cm^-1^) [18,22]. Furthermore, two of these studies utilized the first derivative of IR spectra [17,22]. In our study, the IR spectra among control, OA, and RA groups were similar, with only minor fluctuations in absorbance values. However, our mean spectra (both normalized and first derivative) across groups were similar to the previously reported data [17,18,22]. Moreover, unlike previous ATR-FTIR studies, we did not find statistically significant wavenumbers that could serve as spectral markers or aid in feature reduction [18,22]. Our results suggest that using normalized data from the entire spectral region results in the most generalizable model. Furthermore, derivative spectra did not add value to our model compared to studies by Durlik-Popińska et al. [22] and Lechowicz et al. [17], in which the first derivative had the most value for classification.

The highest model performance between OA and RA groups suggests that the serum biochemical responses differ most between rheumatic diseases. This can also be because OA phenotypes include more biomechanical and structural changes, while RA has metabolic primarily and inflammatory phenotypes. This hypothesis is supported by a previous FTIR study by Wu X et al., which shows the most prominent differences between mean FTIR spectra of ankylosing spondylitis (AS), RA, and OA patients [21]. Moreover, a similar study used deep learning to classify FTIR spectra obtained from AS and RA patients with encouraging results. This also could explain the classification performance between the control RA group since RA is a well-established inflammatory disease with several serological effects. We found the lowest performance between the control and OA groups. This finding may reflect that knee OA’s pathophysiology is mainly due to biomechanical forces [40], and thus, biochemical processes behind knee OA are better reflected in the synovial fluid than in the serum, as systematic effects may play a minor role and are undetectable [41].

We used a nested CV to train the classifier models, which is shown to be the most relatable of true error [34,42,43]. This may yield lower overall performance for the models. A previous study used discriminant analysis as a feature selection technique for all the data before the actual modeling process [18]. This process causes potential data leakage and may lead to overfitting. They also used more than one spectrum from the same individual in modeling. This may lead to an over-optimistic model, primarily when leave-one-patient-out cross-validation (LOPOCV) was not used [18]. Correspondingly, the study by Lechowicz et al.[17] used three spectra replicates from each individual in the KNN model without LOPOCV, uncertainly estimating the model’s generalization ability.

The PLS-DA-SVM method combines aspects of principal component analysis (PCA), linear discriminant analysis (LDA), and canonical correlation analysis (CCA) to reduce the dimensionality of the data [44]. In PLS-DA (or PLS regression), the covariance between predictive and responsible variables is maximized to maximize the difference between sample groups in LV space (PLS-score matrix) [44], in which the response variable information is needed. Therefore, it is essential not to imply this latent LV construction straightaway to the whole data set before training the model due to the evident data leakage and to produce a non-biased, reliable generalization ability estimation of the model. In this study, we acknowledged the challenge of potential over-optimism in model evaluation by applying PLS as a dimension-reduction technique. This approach is the first to handle generalization issues in spectroscopy-based cancer diagnostics effectively [28–31,45,46], alprazolam qualification [27], and the classification of ionic liquids, coffee variants, and olive oils [47]. The latent variable spaces, derived from the equation presented in Section 2.5, demonstrate a more realistic but limited generalization, highlighting a significant step forward for future research.

One limitation of this study is the relatively small sample size and lack of diversity. Additionally, not filtering out abundant serum proteins may have affected the spectra. For instance, Hackshaw et al. utilized a centrifugal membrane filter to eliminate nominal molecular mass blood components in metabolic fingerprinting of Raman spectra of rheumatic disorders [19]. Detecting crucial protein markers can be difficult due to the high levels of proteins in the serum, which might interfere with the spectral signal. Nevertheless, we maintained a consistent measurement setup for all serum samples for comparison and classification. Furthermore, it is crucial to avoid algorithmic bias and acquire sufficient data. However, achieving adequate laboratory data collection requires substantial resources, rendering it a challenging task in the future. The differences in results compared to previous ATR-FTIR studies may also indicate the challenges of ATR-FTIR’s ability to apply to various populations and spectrometers when identifying OA and RA from serum. Our sample set displayed a significant degree of similarity in their biochemical composition, which may have contributed to the consistency of the results. Despite these challenges, the primary strength of our study lies in the use of robust and generalized PLS-DA-SVM classification models.

## 5. Conclusions

In conclusion, our PLS-DA-SVM classifier models performed best while separating OA from RA serum samples. These results indicate that ATR-FTIR has the potential to detect systematic inflammation-induced differences within the human serum. It may be a rapid and economic aid in classifying rheumatic diseases.

## Author contributions: CRediT

Conceptualization: MM, SDG, LR & SS, Data curation: MM, Formal analysis: MM, Investigation: MM, Methodology: MM, Resources: MM & SDG, Supervision: SDG & SS, Writing – original draft: MM & SDG, Writing – review and editing: SDG, LR & SS

## Data Availability

All data produced in the present study are available upon reasonable request to the authors.

## Acknowledgments

We want to acknowledge the clinical staff at Biobank Borealis, Oulu, Finland (https://oys.fi/biopankki/) and sample donors for providing the blood serum sample set.

## References

1. P.H. Hsieh, O. Wu, C. Geue, E. McIntosh, I.B. McInnes, S. Siebert, Economic burden of rheumatoid arthritis: A systematic review of literature in biologic era, Ann Rheum Dis 79 (2020). 10.1136/annrheumdis-2019-216243.

[2] V.P. Leifer, J.N. Katz, E. Losina, The burden of OA-health services and economics, Osteoarthritis Cartilage 30 (2022) 10–16. 10.1016/J.JOCA.2021.05.007.

[3] J.D. Steinmetz, G.T. Culbreth, L.M. Haile, Q. Rafferty, J. Lo, K.G. Fukutaki, J.A. Cruz, A.E. Smith, S.E. Vollset, P.M. Brooks, M. Cross, A.D. Woolf, H. Hagins, M. Abbasi-Kangevari, A. Abedi, I.N. Ackerman, H. Amu, B. Antony, J. Arabloo, A.Y. Aravkin, A.M. Argaw, A.A. Artamonov, T. Ashraf, A. Barrow, L.M. Bearne, I.M. Bensenor, A.Y. Berhie, N. Bhardwaj, P. Bhardwaj, V.S. Bhojaraja, A. Bijani, P.S. Briant, A.M. Briggs, N.S. Butt, J. Charan, V.K. Chattu, F.M. Cicuttini, K. Coberly, O. Dadras, X. Dai, L. Dandona, R. Dandona, K. de Luca, E. Denova-Gutiérrez, S.D. Dharmaratne, M. Dhimal, M. Dianatinasab, K.E. Dreinhoefer, M. Elhadi, U. Farooque, H.R. Farpour, I. Filip, F. Fischer, M. Freitas, B. Ganesan, B.N.B. Gemeda, T. Getachew, S.H. Ghamari, A. Ghashghaee, T.K. Gill, M. Golechha, D. Golinelli, B. Gupta, V.B. Gupta, V.K. Gupta, R. Haddadi, N. Hafezi-Nejad, R. Halwani, S. Hamidi, A. Hanif, N.I. Harlianto, J.M. Haro, J. Hartvigsen, S.I. Hay, J.J. Hebert, G. Heidari, M.S. Hosseini, M. Hosseinzadeh, A.K. Hsiao, I.M. Ilic, M.D. Ilic, L. Jacob, R. Jayawardena, R.P. Jha, J.B. Jonas, N. Joseph, H. Kandel, I.M. Karaye, M.J. Khan, Y.J. Kim, A.A. Kolahi, O. Korzh, R. Koteeswaran, V. Krishnamoorthy, G.A. Kumar, N. Kumar, S. woong Lee, S.S. Lim, S.W. Lobo, G. Lucchetti, M.R. Malekpour, A.A. Malik, L.G.G. Mandarano-Filho, S. Martini, A.F.A. Mentis, M.K. Mesregah, T. Mestrovic, E.M. Mirrakhimov, A. Misganaw, R. Mohammadpourhodki, A.H. Mokdad, S. Momtazmanesh, S.D. Morrison, C.J.L. Murray, H. Nassereldine, H.B. Netsere, S. Neupane Kandel, M.O. Owolabi, S. Panda-Jonas, A. Pandey, S. Pawar, P. Pedersini, J. Pereira, A. Radfar, M.M. Rashidi, D.L. Rawaf, S. Rawaf, R. Rawassizadeh, S.M. Rayegani, D. Ribeiro, L. Roever, B. Saddik, A. Sahebkar, S. Salehi, L. Sanchez Riera, F. Sanmarchi, M.M. Santric-Milicevic, S. Shahabi, M.A. Shaikh, E. Shaker, M. Shannawaz, R. Sharma, S. Sharma, J.K. Shetty, R. Shiri, P. Shobeiri, D.A.S. Silva, A. Singh, J.A. Singh, S. Singh, S.T. Skou, H. Slater, M.S. Soltani-Zangbar, A. V. Starodubova, A. Tehrani-Banihashemi, S. Valadan Tahbaz, P.R. Valdez, B. Vo, L.G. Vu, Y.P. Wang, S.H. Yahyazadeh Jabbari, N. Yonemoto, I. Yunusa, L.M. March, K.L. Ong, T. Vos, J.A. Kopec, Global, regional, and national burden of osteoarthritis, 1990–2020 and projections to 2050: a systematic analysis for the Global Burden of Disease Study 2021, Lancet Rheumatol 5 (2023). 10.1016/S2665-9913(23)00163-7.

[4] D.J. Hunter, S. Bierma-Zeinstra, Osteoarthritis, The Lancet 393 (2019) 1745–1759. 10.1016/S0140-6736(19)30417-9.

[5] E.M. Gravallese, G.S. Firestein, Rheumatoid Arthritis — Common Origins, Divergent Mechanisms, New England Journal of Medicine 388 (2023). 10.1056/nejmra2103726.

[6] A. Di Matteo, J.M. Bathon, P. Emery, Rheumatoid arthritis, The Lancet 402 (2023) 2019– 2033. 10.1016/S0140-6736(23)01525-8.

[7] V.B. Kraus, F.J. Blanco, M. Englund, M.A. Karsdal, L.S. Lohmander, Call for standardized definitions of osteoarthritis and risk stratification for clinical trials and clinical use, Osteoarthritis Cartilage 23 (2015) 1233–1241. 10.1016/J.JOCA.2015.03.036.

[8] D. Aletaha, T. Neogi, A.J. Silman, J. Funovits, D.T. Felson, C.O. Bingham, N.S. Birnbaum, G.R. Burmester, V.P. Bykerk, M.D. Cohen, B. Combe, K.H. Costenbader, M. Dougados, P. Emery, G. Ferraccioli, J.M.W. Hazes, K. Hobbs, T.W.J. Huizinga, A. Kavanaugh, J. Kay, T.K. Kvien, T. Laing, P. Mease, H.A. Ménard, L.W. Moreland, R.L. Naden, T. Pincus, J.S. Smolen, E. Stanislawska-Biernat, D. Symmons, P.P. Tak, K.S. Upchurch, J. Vencovský, F. Wolfe, G. Hawker, 2010 Rheumatoid arthritis classification criteria: An American College of Rheumatology/European League Against Rheumatism collaborative initiative, Arthritis Rheum 62 (2010) 2569–2581. 10.1002/art.27584.

[9] NICE, Osteoarthritis in over 16s: diagnosis and management, NICE Guideline (2022).

[10] Q. Wang, J. Runhaar, M. Kloppenburg, M. Boers, J.W.J. Bijlsma, S.M.A. Bierma-Zeinstra, Evaluation of the Diagnostic Performance of American College of Rheumatology, EULAR, and National Institute for Health and Clinical Excellence Criteria Against Clinically Relevant Knee Osteoarthritis: Data From the CHECK Cohort, Arthritis Care Res (Hoboken) (2023). 10.1002/acr.25270.

11. H. Radner, T. Neogi, J.S. Smolen, D. Aletaha, Performance of the 2010 ACR/EULAR classification criteria for rheumatoid arthritis: A systematic literature review, Ann Rheum Dis 73 (2014). 10.1136/annrheumdis-2013-203284.

[12] H.J. Butler, P.M. Brennan, J.M. Cameron, D. Finlayson, M.G. Hegarty, M.D. Jenkinson, D.S. Palmer, B.R. Smith, M.J. Baker, Development of high-throughput ATR-FTIR technology for rapid triage of brain cancer, Nat Commun 10 (2019). 10.1038/s41467-019-12527-5.

[13] S. Roy, D. Perez-Guaita, S. Bowden, P. Heraud, B.R. Wood, Spectroscopy goes viral: Diagnosis of hepatitis B and C virus infection from human sera using ATR-FTIR spectroscopy, Clinical Spectroscopy 1 (2019). 10.1016/j.clispe.2020.100001.

[14] S. Ali, A. Naveed, I. Hussain, J. Qazi, Use of ATR-FTIR spectroscopy to differentiate between cirrhotic/non-cirrhotic HCV patients, Photodiagnosis Photodyn Ther 42 (2023). 10.1016/j.pdpdt.2023.103529.

[15] V.E. Sitnikova, M.A. Kotkova, T.N. Nosenko, T.N. Kotkova, D.M. Martynova, M. V. Uspenskaya, Breast cancer detection by ATR-FTIR spectroscopy of blood serum and multivariate data-analysis, Talanta 214 (2020). 10.1016/j.talanta.2020.120857.

[16] P.J. Larkin, Instrumentation and Sampling Methods, Infrared and Raman Spectroscopy (2018) 29–61. 10.1016/B978-0-12-804162-8.00003-3.

[17] L. Lechowicz, M. Chrapek, J. Gaweda, M. Urbaniak, I. Konieczna, Use of Fourier-transform infrared spectroscopy in the diagnosis of rheumatoid arthritis: a pilot study, Mol Biol Rep 43 (2016) 1321–1326. 10.1007/s11033-016-4079-7.

[18] A.A. Aziz, V. Selvaratnam, Y.F.B.A. Fikri, M.S.A. Sani, T. Kamarul, Diagnosis of Osteoarthritis at an Early Stage via Infrared Spectroscopy Combined Chemometrics in Human Serum: A Pilot Study, Processes 11 (2023). 10.3390/pr11020404.

[19] K. V. Hackshaw, D.P. Aykas, G.T. Sigurdson, M. Plans, F. Madiai, L. Yu, C.A.T. Buffington, M. Mónica Giusti, L. Rodriguez-Saona, Metabolic fingerprinting for diagnosis of fibromyalgia and other rheumatologic disorders, Journal of Biological Chemistry 294 (2019) 2555–2568. 10.1074/jbc.RA118.005816.

[20] W. Shuai, X. Wu, C. Chen, E. Zuo, X. Chen, Z. Li, X. Lv, L. Wu, C. Chen, Rapid diagnosis of rheumatoid arthritis and ankylosing spondylitis based on Fourier transform infrared spectroscopy and deep learning, Photodiagnosis Photodyn Ther 45 (2024) 103885. 10.1016/J.PDPDT.2023.103885.

[21] X. Wu, W. Shuai, C. Chen, X. Chen, C. Luo, Y. Chen, Y. Shi, Z. Li, X. Lv, C. Chen, X. Meng, X. Lei, L. Wu, Rapid screening for autoimmune diseases using Fourier transform infrared spectroscopy and deep learning algorithms, Front Immunol 14 (2023). 10.3389/fimmu.2023.1328228.

[22] K. Durlik-Popińska, P. Żarnowiec, I. Konieczna-Kwinkowska, Ł. Lechowicz, J. Gawęda, W. Kaca, Correlations between autoantibodies and the ATR-FTIR spectra of sera from rheumatoid arthritis patients, Sci Rep 11 (2021). 10.1038/s41598-021-96848-w.

[23] H.H. Eysel, M. Jackson, A. Nikulin, R.L. Somorjai, G.T.D. Thomson, H.H. Mantsch, A novel diagnostic test for arthritis: Multivariate analysis of infrared spectra of synovial fluid, Biospectroscopy 3 (1997). 10.1002/(SICI)1520-6343(1997)3:2<161::AID-BSPY9>3.0.CO;2-A.

[24] P. Galozzi, D. Basso, M. Plebani, A. Padoan, Artificial intelligence and laboratory data in rheumatic diseases, Clinica Chimica Acta 546 (2023) 117388. 10.1016/J.CCA.2023.117388.

[25] L.B. Leal, M.S. Nogueira, R.A. Canevari, L.F.C.S. Carvalho, Vibration spectroscopy and body biofluids: Literature review for clinical applications, Photodiagnosis Photodyn Ther 24 (2018) 237–244. 10.1016/J.PDPDT.2018.09.008.

[26] A. Aït-Kaddour, M. Loudiyi, A. Ferlay, D. Gruffat, Performance of fluorescence spectroscopy for beef meat authentication: Effect of excitation mode and discriminant algorithms, Meat Sci 137 (2018) 58–66. 10.1016/J.MEATSCI.2017.11.002.

[27] E. Parhizkar, M. Ghazali, F. Ahmadi, A. Sakhteman, PLS-LS-SVM based modeling of ATR-IR as a robust method in detection and qualification of alprazolam, Spectrochim Acta A Mol Biomol Spectrosc 173 (2017) 87–92. 10.1016/J.SAA.2016.08.055.

[28] T. Sun, Y. Lin, Y. Yu, S. Gao, X. Gao, H. Zhang, K. Lin, J. Lin, Low-abundance proteins-based label-free SERS approach for high precision detection of liver cancer with different stages, Anal Chim Acta 1304 (2024) 342518. 10.1016/J.ACA.2024.342518.

[29] Y. Lin, J. Gao, S. Tang, X. Zhao, M. Zheng, W. Gong, S. Xie, S. Gao, Y. Yu, J. Lin, Label-free diagnosis of breast cancer based on serum protein purification assisted surface-enhanced Raman spectroscopy, Spectrochim Acta A Mol Biomol Spectrosc 263 (2021) 120234. 10.1016/J.SAA.2021.120234.

[30] X. Bai, J. Lin, X. Wu, Y. Lin, X. Zhao, W. Du, J. Gao, Z. Hu, Q. Xu, T. Li, Y. Yu, Label-free detection of bladder cancer and kidney cancer plasma based on SERS and multivariate statistical algorithm, Spectrochim Acta A Mol Biomol Spectrosc 279 (2022) 121336. 10.1016/J.SAA.2022.121336.

[31] S. Gao, Y. Lin, X. Zhao, J. Gao, S. Xie, W. Gong, Y. Yu, J. Lin, Label-free surface enhanced Raman spectroscopy analysis of blood serum via coffee ring effect for accurate diagnosis of cancers, Spectrochim Acta A Mol Biomol Spectrosc 267 (2022) 120605. 10.1016/J.SAA.2021.120605.

[32] S. Elderderi, C. Leman-Loubière, L. Wils, S. Henry, D. Bertrand, H.J. Byrne, I. Chourpa, C. Enguehard-Gueiffier, E. Munnier, A.A. Elbashir, L. Boudesocque-Delaye, F. Bonnier, ATR-IR spectroscopy for rapid quantification of water content in deep eutectic solvents, J Mol Liq 311 (2020) 113361. 10.1016/J.MOLLIQ.2020.113361.

[33] C.L.M. Morais, M. Paraskevaidi, L. Cui, N.J. Fullwood, M. Isabelle, K.M.G. Lima, P.L. Martin-Hirsch, H. Sreedhar, J. Trevisan, M.J. Walsh, D. Zhang, Y.G. Zhu, F.L. Martin, Standardization of complex biologically derived spectrochemical datasets, Nat Protoc 14 (2019). 10.1038/s41596-019-0150-x.

[34] B. Bischl, M. Binder, M. Lang, T. Pielok, J. Richter, S. Coors, J. Thomas, T. Ullmann, M. Becker, A.L. Boulesteix, D. Deng, M. Lindauer, Hyperparameter optimization: Foundations, algorithms, best practices, and open challenges, Wiley Interdiscip Rev Data Min Knowl Discov 13 (2023). 10.1002/widm.1484.

[35] A.C.S. Talari, M.A.G. Martinez, Z. Movasaghi, S. Rehman, I.U. Rehman, Advances in Fourier transform infrared (FTIR) spectroscopy of biological tissues, Appl Spectrosc Rev 52 (2017). 10.1080/05704928.2016.1230863.

[36] S. Olsztyńska-Janus, K. Szymborska-Małek, M. Gasior-Glogowska, T. Walski, M. Komorowska, W. Witkiewicz, C. Pezowicz, M. Kobielarz, S. Szotek, Spectroscopic techniques in the study of human tissues and their components. Part I: IR spectroscopy, Acta Bioeng Biomech 14 (2012). 10.5277/abb120314.

[37] N. Oskam, P. Ooijevaar-De Heer, D. Kos, J. Jeremiasse, L. Van Boheemen, G.M. Verstappen, F.G.M. Kroese, D. Van Schaardenburg, G. Wolbink, T. Rispens, Rheumatoid factor autoantibody repertoire profiling reveals distinct binding epitopes in health and autoimmunity, Ann Rheum Dis 82 (2023). 10.1136/ard-2023-223901.

[38] L. Voronina, F. Fleischmann, J. Šimunović, C. Ludwig, M. Novokmet, M. Žigman, Probing Blood Plasma Protein Glycosylation with Infrared Spectroscopy, Anal Chem (2023). 10.1021/acs.analchem.3c03589.

[39] D. Sun, F. Hu, H. Gao, Z. Song, W. Xie, P. Wang, L. Shi, K. Wang, Y. Li, C. Huang, Z. Li, Distribution of abnormal IgG glycosylation patterns from rheumatoid arthritis and osteoarthritis patients by MALDI-TOF-MS:N, Analyst 144 (2019). 10.1039/c8an02014k.

[40] M. Englund, The role of biomechanics in the initiation and progression of OA of the knee, Best Pract Res Clin Rheumatol 24 (2010) 39–46. 10.1016/J.BERH.2009.08.008.

[41] A. Mahmoudian, L.S. Lohmander, A. Mobasheri, M. Englund, F.P. Luyten, Early-stage symptomatic osteoarthritis of the knee — time for action, Nat Rev Rheumatol 17 (2021). 10.1038/s41584-021-00673-4.

[42] S. Bates, T. Hastie, R. Tibshirani, Cross-Validation: What Does It Estimate and How Well Does It Do It?, J Am Stat Assoc 119 (2024). 10.1080/01621459.2023.2197686.

[43] S. Varma, R. Simon, Bias in error estimation when using cross-validation for model selection, BMC Bioinformatics 7 (2006). 10.1186/1471-2105-7-91.

[44] S. de Jong, SIMPLS: An alternative approach to partial least squares regression, Chemometrics and Intelligent Laboratory Systems 18 (1993) 251–263. 10.1016/0169-7439(93)85002-X.

[45] X. Li, T. Yang, S. Li, L. Jin, D. Wang, D. Guan, J. Ding, Noninvasive liver diseases detection based on serum surface enhanced Raman spectroscopy and statistical analysis, Opt Express 23 (2015). 10.1364/oe.23.018361.

[46] S. Gao, M. Zheng, Y. Lin, K. Lin, J. Zeng, S. Xie, Y. Yu, J. Lin, Surface-enhanced Raman scattering analysis of serum albumin via adsorption-exfoliation on hydroxyapatite nanoparticles for noninvasive cancers screening, J Biophotonics 13 (2020). 10.1002/jbio.202000087.

[47] A. Kanwal, T. Mehmood, M.M. Butt, PLS and kernel SVM based hybrid classifier for discriminating FTIR spectrum data with limited sample size, Chemometrics and Intelligent Laboratory Systems 215 (2021) 104365. 10.1016/J.CHEMOLAB.2021.104365.

