## Supplementary for "Diagnostic performance of Attenuated Total Reflectance Fourier Transform Infrared (ATR-FTIR) spectroscopy for detecting osteoarthritis and rheumatoid arthritis from blood serum"

<u>**Electronic Supplemental Material**</u>

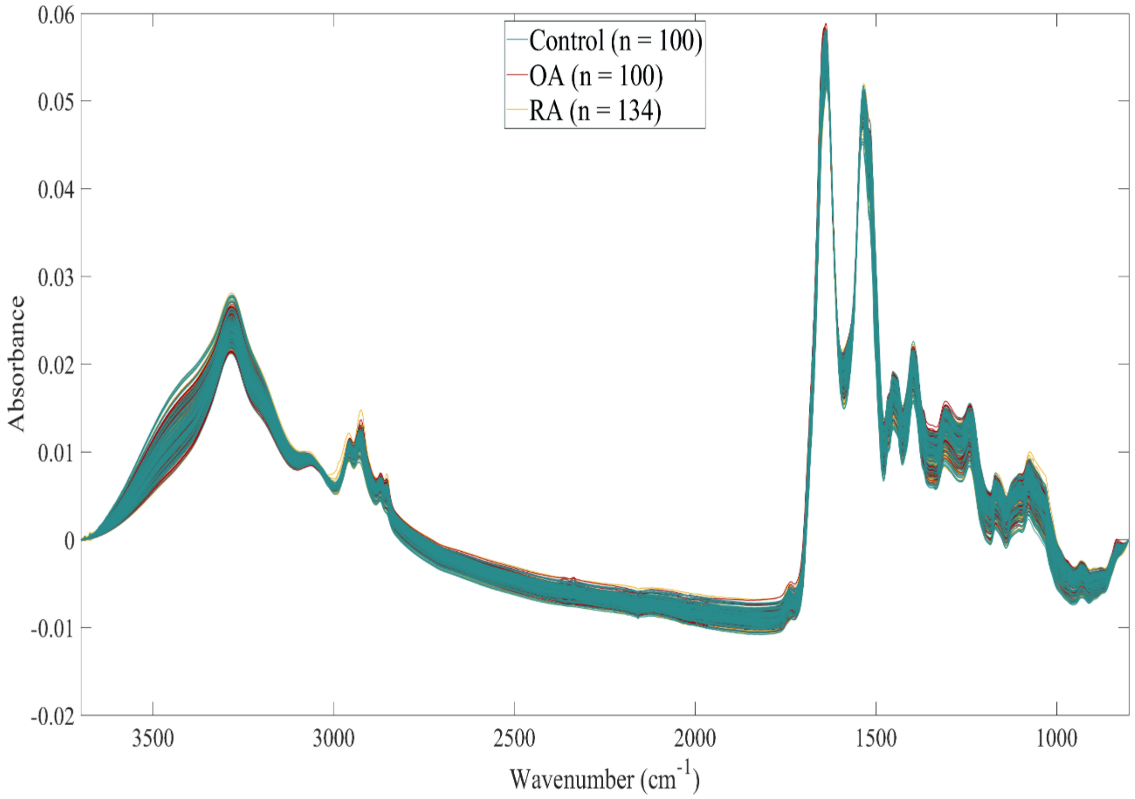


**Figure S1.** Illustration of all 334 preprocessed ATR-FTIR spectra. Bluish-green corresponds to all spectra in the control group, red in the OA group, and yellow in the RA group.

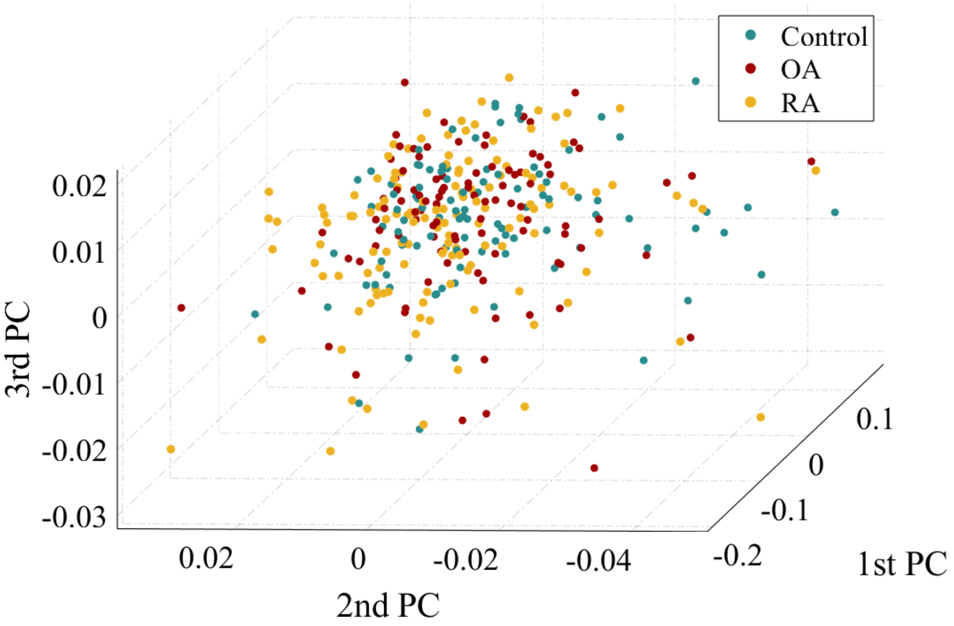


**Figure S2**. The first three principal components of preprocessed spectra truncated to the 800 – 3700 $cm^{-1}$ wavenumber region. The figure shows that PCA cannot detect discriminative patterns between groups from the spectral data, making it an inadequate dimension reduction method before classification.

Figures S1-S2 show that the sample-to-sample variations are higher than the subtle differences between groups, which led to PCA's inability to distinguish discriminative patterns between groups.

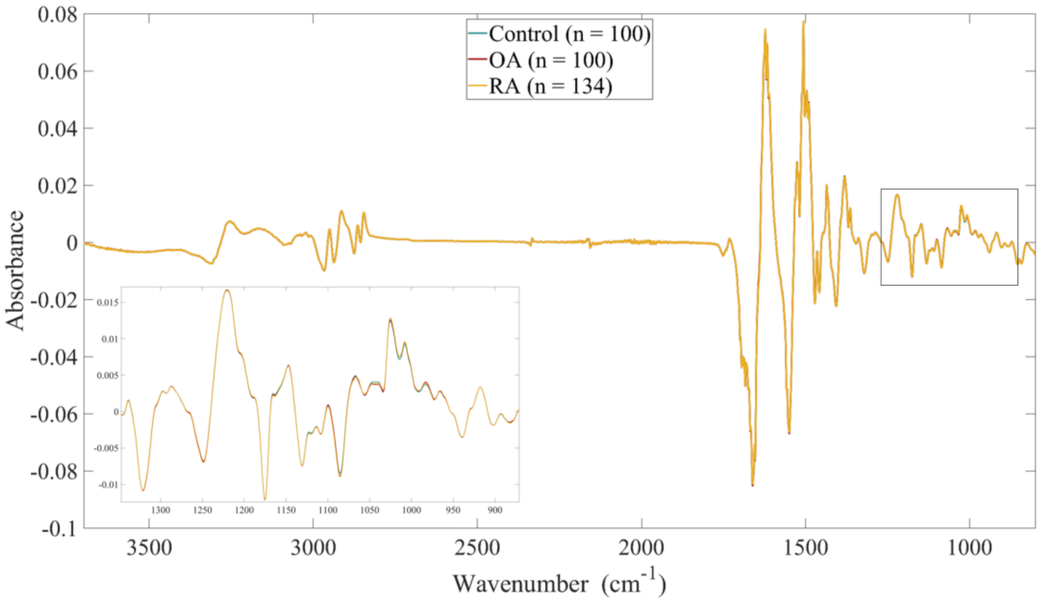


**Figure S3.** Illustration of the mean first derivative spectra, truncated into the 800 – 3700 cm$^{-1}$ wavenumber region. The blue spectrum represents the mean spectrum of the control group, the red spectrum represents the mean spectrum of the OA group, and the yellow spectrum represents the mean spectrum of the RA group. As it is clear, no significant differences are shown between groups. The black rectangle highlights the minimal absorbance differences observed in the lower wavenumbers (900 – 1400 cm$^{-1}$).

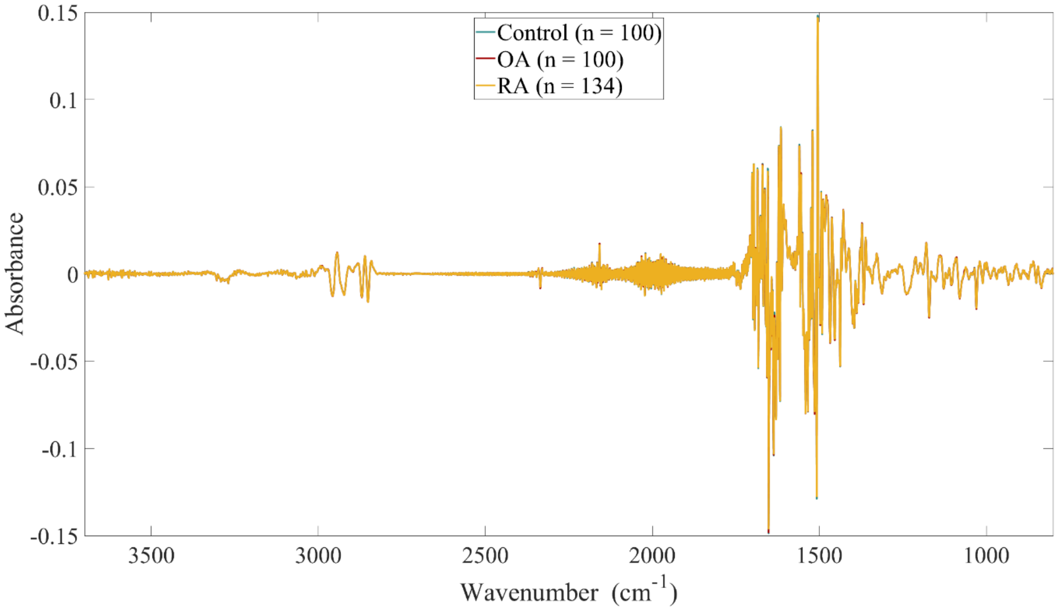


**Figure S4.** Illustration of the mean second derivative spectra, truncated into the 800 – 3700 cm$^{-1}$ wavenumber region. The blue spectrum represents the mean spectrum of the control group, the red spectrum represents the mean spectrum of the OA group, and the yellow spectrum represents the mean spectrum of the RA group. As it is clear, no significant differences are shown between groups.

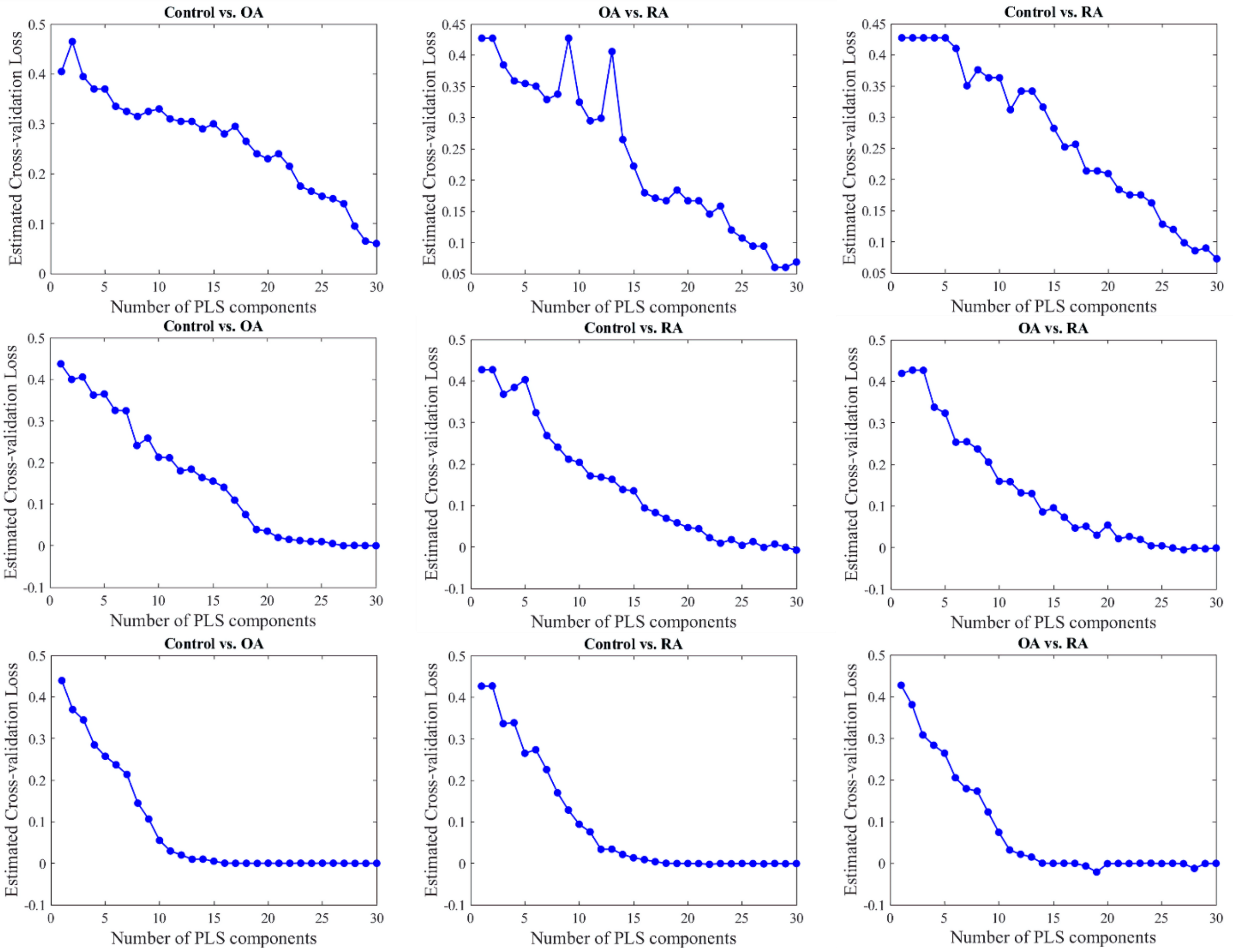


**Figure S5.** The estimated cross-validation errors for SVM. The first row corresponds to the three binary classifications performed with normalized spectral data, the second to the first derivative spectral data, and the third to the second derivative spectral data. The spectra were truncated into 800 – 3700 $cm^{-1}$, wavenumber region.

**Table S1.** The PLS-SVM classifier's highest-observed performance on validation and test sets was when normalized spectral data truncated into the 800 – 1800 $cm^{-1}$ wavenumber region was used.

| Performance metrics | | AUC-ROC | Accuracy (%) | Sensitivity (%) | Specificity (%) | Number of PLS-components |
|---|---|---|---|---|---|---|
| **Validation set** | Control vs. OA | 0.76 | 72 | 67 | 74 | 10 |
| | Control vs. RA | 0.88 | 80 | 83 | 78 | 20 |
| | OA vs. RA | 0.92 | 86 | 92 | 78 | 20 |
| **Test set** | Control vs. OA | 0.70 | 60 | 61 | 59 | 10 |
| | Control vs. RA | 0.76 | 80 | 90 | 66 | 20 |
| | OA vs. RA | 0.85 | 81 | 86 | 74 | 20 |

**Table S2.** The averaged performance values on the validation and test set aggregated across the 100 iterations of PLS-SVM model training when normalized spectral data, truncated into the 800 – 1800 $cm^{-1}$ wavenumber region, was used.

| Performance metrics | | AUC-ROC | Accuracy (%) | Sensitivity (%) | Specificity (%) | Number of PLS-components |
|---|---|---|---|---|---|---|
| **Validation set** | Control vs. OA | 0.66 | 62 | 61 | 65 | 10 |
| | Control vs. RA | 0.83 | 76 | 82 | 69 | 20 |
| | OA vs. RA | 0.87 | 81 | 85 | 76 | 20 |
| **Test set** | Control vs. OA | 0.55 | 53 | 52 | 54 | 10 |
| | Control vs. RA | 0.64 | 61 | 38 | 93 | 20 |
| | OA vs. RA | 0.72 | 66 | 45 | 95 | 20 |

**Table S3.** The PLS-SVM classifier's highest-observed performance on validation and test sets was when the first derivative spectral data truncated into the 800 – 1800 cm$^{-1}$ wavenumber region was used.

| **Performance metrics** | | **AUC-ROC** | **Accuracy (%)** | **Sensitivity (%)** | **Specificity (%)** | **Number of PLS-components** |
|---|---|---|---|---|---|---|
| **Validation set** | Control vs. OA | 0.86 | 81 | 78 | 87 | 10 |
| | Control vs. RA | 0.97 | 93 | 95 | 90 | 15 |
| | OA vs. RA | 0.99 | 99 | 99 | 100 | 20 |
| **Test set** | Control vs. OA | 0.70 | 58 | 54 | 63 | 10 |
| | Control vs. RA | 0.76 | 63 | 60 | 65 | 15 |
| | OA vs. RA | 0.81 | 79 | 79 | 78 | 20 |

**Table S4.** The averaged performance values on the validation and test set aggregated across the 100 iterations of PLS-SVM model training when first derivative spectral data, truncated into the 800 – 1800 cm$^{-1}$ wavenumber region, was used.

| **Performance metrics** | | **AUC-ROC** | **Accuracy (%)** | **Sensitivity (%)** | **Specificity (%)** | **Number of PLS-components** |
|---|---|---|---|---|---|---|
| **Validation set** | Control vs. OA | 0.79 | 73 | 71 | 75 | 10 |
| | Control vs. RA | 0.93 | 86 | 89 | 85 | 15 |
| | OA vs. RA | 0.99 | 97 | 97 | 96 | 20 |
| **Test set** | Control vs. OA | 0.58 | 55 | 54 | 58 | 10 |
| | Control vs. RA | 0.64 | 59 | 38 | 90 | 15 |
| | OA vs. RA | 0.70 | 72 | 63 | 75 | 20 |

**Table S5.** The PLS-SVM classifier's highest-observed performance on validation and test sets was when second derivative spectral data truncated into the 800 – 1800 $cm^{-1}$ wavenumber region was used.

| Performance metrics | | AUC-ROC | Accuracy (%) | Sensitivity (%) | Specificity (%) | Number of PLS-components |
|---|---|---|---|---|---|---|
| Validation set | Control vs. OA | 0.79 | 76 | 79 | 70 | 5 |
| | Control vs. RA | 0.76 | 74 | 69 | 80 | 5 |
| | OA vs. RA | 1 | 100 | 100 | 100 | 20 |
| Test set | Control vs. OA | 0.72 | 50 | 0 | 100 | 5 |
| | Control vs. RA | 0.69 | 66 | 81 | 58 | 5 |
| | OA vs. RA | 0.74 | 71 | 81 | 61 | 20 |

**Table S6.** The averaged performance values on the validation and test set aggregated across the 100 iterations of PLS-SVM model training when second derivative spectral data, truncated into the 800 – 1800 $cm^{-1}$ wavenumber region, was used.

| Performance metrics | | AUC-ROC | Accuracy (%) | Sensitivity (%) | Specificity (%) | Number of PLS-components |
|---|---|---|---|---|---|---|
| Validation set | Control vs. OA | 0.72 | 66 | 66 | 67 | 5 |
| | Control vs. RA | 0.68 | 67 | 73 | 59 | 5 |
| | OA vs. RA | 1 | 100 | 100 | 100 | 20 |
| Test set | Control vs. OA | 0.55 | 52 | 53 | 52 | 5 |
| | Control vs. RA | 0.56 | 62 | 78 | 38 | 5 |
| | OA vs. RA | 0.56 | 62 | 40 | 75 | 20 |

**Table S7**. The PLS-SVM classifier's highest-observed performance on validation and test sets was when the first derivative spectral data truncated into the 800 – 3700 cm$^{-1}$ wavenumber region was used.

| **Performance metrics** | | **AUC-ROC** | **Accuracy (%)** | **Sensitivity (%)** | **Specificity (%)** | **Number of PLS-components** |
|---|---|---|---|---|---|---|
| **Validation set** | Control vs. OA | 0.99 | 99 | 97 | 100 | 10 |
| | Control vs. RA | 0.99 | 96 | 93 | 100 | 10 |
| | OA vs. RA | 0.99 | 96 | 97 | 96 | 10 |
| **Test set** | Control vs. OA | 0.71 | 65 | 69 | 63 | 10 |
| | Control vs. RA | 0.68 | 70 | 79 | 56 | 10 |
| | OA vs. RA | 0.82 | 77 | 84 | 69 | 10 |

**Table S8.** The averaged performance values on the validation and test set aggregated across the 100 iterations of PLS-SVM model training when the first derivative data, truncated into the 800 – 3700 cm$^{-1}$ wavenumber region, was used.

| **Performance metrics** | | **AUC-ROC** | **Accuracy (%)** | **Sensitivity (%)** | **Specificity (%)** | **Number of PLS-components** |
|---|---|---|---|---|---|---|
| **Validation set** | Control vs. OA | 0.98 | 94 | 93 | 95 | 10 |
| | Control vs. RA | 0.97 | 92 | 91 | 93 | 10 |
| | OA vs. RA | 0.97 | 93 | 96 | 90 | 10 |
| **Test set** | Control vs. OA | 0.55 | 61 | 56 | 62 | 10 |
| | Control vs. RA | 0.56 | 62 | 80 | 33 | 10 |
| | OA vs. RA | 0.68 | 68 | 69 | 64 | 10 |

**Table S9.** The PLS-SVM classifier's highest-observed performance on validation and test sets was when second derivative spectral data truncated into the 800 – 3700 $cm^{-1}$ wavenumber region was used.

| **Performance metrics** | | **AUC-ROC** | **Accuracy (%)** | **Sensitivity (%)** | **Specificity (%)** | **Number of PLS-components** |
|---|---|---|---|---|---|---|
| **Validation set** | Control vs. OA | 1 | 100 | 100 | 100 | 10 |
| | Control vs. RA | 0.98 | 93 | 91 | 94 | 10 |
| | OA vs. RA | 0.96 | 88 | 85 | 90 | 5 |
| **Test set** | Control vs. OA | 0.60 | 63 | 64 | 63 | 10 |
| | Control vs. RA | 0.70 | 74 | 74 | 75 | 10 |
| | OA vs. RA | 0.72 | 74 | 71 | 78 | 5 |

**Table S10.** The averaged performance values on the validation and test set aggregated across the 100 iterations of PLS-SVM model training when second derivative spectral data, truncated into the 800 – 3700 $cm^{-1}$ wavenumber region, was used.

| **Performance metrics** | | **AUC-ROC** | **Accuracy (%)** | **Sensitivity (%)** | **Specificity (%)** | **Number of PLS-components** |
|---|---|---|---|---|---|---|
| **Validation set** | Control vs. OA | 1 | 100 | 100 | 100 | 10 |
| | Control vs. RA | 0.96 | 90 | 90 | 91 | 10 |
| | OA vs. RA | 0.93 | 88 | 90 | 85 | 5 |
| **Test set** | Control vs. OA | 0.55 | 61 | 52 | 68 | 10 |
| | Control vs. RA | 0.58 | 65 | 59 | 64 | 10 |
| | OA vs. RA | 0.59 | 62 | 60 | 54 | 5 |
